# Epidemiological indices with multiple circulating pathogen strains

**DOI:** 10.1101/2024.12.11.24318844

**Authors:** Cristiano Trevisin, Lorenzo Mari, Marino Gatto, Vittoria Colizza, Andrea Rinaldo

## Abstract

Epidemiological indicators, such as reproduction numbers and epidemicity indices, describe an ongoing epidemic’s long- and short-term behaviour. Their evolving values provide context for designing control measures, as maintaining both indices below one warrants a waning epidemic. However, current models for the computation of epidemiological metrics do not consider the pathogen stratification into variants endowed with different infectivity and epidemiological severity, as was the case with SARS-CoV-2. Failing to account for such stratification prevents the resulting epidemiological indices from spotting the possible onset of uncontrolled growth of one variant within the total of new infections, thus limiting the prognostic value of the indicators. Here, we expand an existing framework for the computation of spatially explicit reproduction numbers and epidemicity indices to account for arising variants. By contrasting the data of the COVID-19 pandemic in Italy, we show that embedding additional layers of complexity in the mathematical model reveals epidemiological metrics at the emergence of new variants significantly exceeding their threshold. Such values foresee a recrudescence in new infections that only became evident after emerging variants effectively replaced the previous strains. These higher values flag the presence of a specific variant that grows more rapidly than the growth of the total number of infections generated by all variants combined. Variant-stratified epidemiological indicators do therefore allow control measures to be better tailored to the shifting severity of an ongoing epidemic.

## Introduction

After its emergence in late 2019, the virus responsible for the COVID-19 pandemic, SARS-CoV-2, has undergone many mutations. The major effort that since the beginning of the pandemic has been devoted to stem the spread of the virus (both vaccines and non-pharmaceutical interventions) has generated a strong evolutionary pressure on the virus to evade immunity conferred by prior vaccination and infection [1, 2, 3]. In particular, this occurred by achieving higher reproduction numbers, shorter doubling times [3, 4] and reduced mean generation times [5, 6] shown by emerged strains. This has not only affected SARS-CoV-2 but also other infectious diseases e.g. dengue fever and influenza. The long-term fate of an epidemic is recapitulated by the computation of the effective reproduction number, say ℛ, which measures the number of secondary infections produced by each primary infector within a given community. Values of ℛ above one flag an accelerating epidemic, whereas subunit values characterize a waning epidemic. In practice, reproduction numbers guide stricter or milder restrictions, with control measures aiming to achieve subunit values. Depending on the chosen epidemiological modelling framework, ℛ may be computed via next-generation matrices, whenever a SIR-like compartmental model is adopted [7, 8], or via renewal equations [9] that allow one to compute reproduction numbers from data directly.

Instead, epidemicity indices describe the stability of disease-free equilibria [10, 11, 12, 13], achievable by attaining a ℛ below its epidemic (ℛ = 1) threshold. However, ℛ < 1 does not prevent epidemic outbursts of infections from occurring[13]. It has been demonstrated that mathematically excluding the possibility of coalescing short-term flare-ups requires ℛ being substantially lower than its epidemic threshold [14, 13, 15].

Within a metapopulation, infections may be transmitted among spatially connected communities via, say, contacts driven by human mobility. The COVID-19 pandemic highlighted the relevance of spatial connectivity both in understanding the spread of SARS-CoV-2 and in identifying proper control measures. Spatial formulations of both the reproduction number [16, 17] and the epidemicity index [15] were derived. Results highlighted how incorporating a spatial stratification yields improvements to estimates of ℛ, as they account for exportation and importation of infections among connected communities, thereby allowing a more precise identification of appropriate control measures. Another stratification, that of the emergence of different variants of the virus of interest, is still missing to date. We posit that embedding strain diversification in frame-works for the computation of epidemiological indices shifts the trajectory of the metrics at the onset of successive variants, often characterized by different reproduction numbers and generation times. To do so, we expand an existing spatially explicit model for the computation of epidemiological metrics [13, 15] to provide a comprehensive framework that seamlessly derives, in real-time, both the reproduction number and the epidemicity index in both space and variants. Accounting for the pathogen’s stratification in both space and variants singles out the fastest-growing strain within the total number of new infections and anticipates a possible recrudescence in new infections.

## Methods

We propose a framework based on a Leslie projection matrix already developed in previous studies [13, 15] and here expanded to account for variant stratification. A key assumption is that, once a variant has emerged, it acts independently of the other circulating strains and that any interaction (most notably, competition) among variants is fully subsumed in their respective ℛ and generation time’s distribution (*β*(*τ*) i.e., the period between the infector’s exposure and that of their infectee(s)). Coinfection has also been document for COVID-19, where two variants have simultaneously infected an individual [18, 19]. These infections may be considered separately in our framework, as the individual may be able to spread both variants at the same time.

To this end, let us consider a metapopulation composed of *N* interconnected human communities, where several variant genotypes (*v* = 1*,…, V*) of a generic infectious disease are circulating. The compartment infected with strain *v* in a generic community *j* (*j* = 1*,…, N*) is indicated as 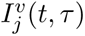, a variable of both time *t* and age of infection *τ*. Also, let *C_lj_*(*t*) be the proportion of residents of the community *j* who commute daily to *l* for their daily activities [16]. The following differential problem holds [16, 13]:

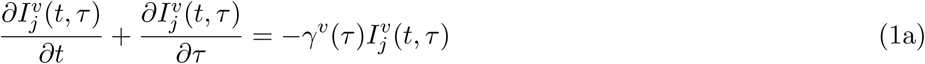

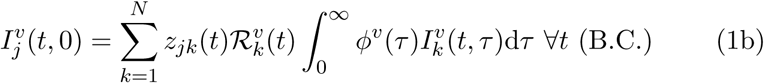

where *γ^v^*(*τ*) represents the instantaneous rate of exit from the infected compartment (because of recovery, death, quarantine, or isolation), and *ϕ^v^*(*τ*) is the rate of secondary transmission per single infectious case. Regardless of the variant being considered, elements *z_jk_* are defined as

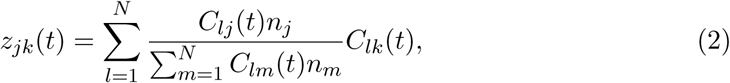

and denote each node’s exposure to infectious individuals residing in a different one via several routes of infection, e.g., individuals living in *j* being exposed to infectors originating in the community *k*, regardless of the community of actual exposure *l* (where two or all *l, k, j* may coincide). Both matrices **C** (of terms *C_lj_*) and **Z** (of terms *z_lj_*) are defined as column-stochastic matrices, with a column-wise sum equalling 1. The rate of secondary transmission *ϕ^v^*(*τ*) relates to the distribution of generation times, *β^v^*(*τ*), via the following relation:

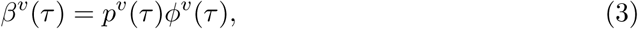

where 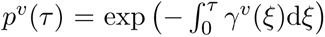 is the survival function, e.g., the fraction of infected individuals that are still infectious after *τ* days. To comply with a daily or weekly collection of epidemiological data, such as the newly detected infection, one may discretize the above process with respect to both the time and the age of infection.

One way of discretizing process (1) leads to the following system:

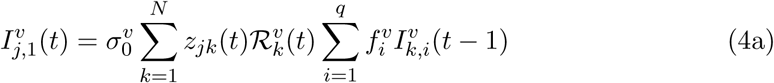

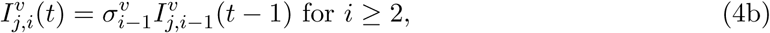

where 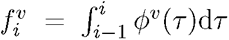 is a discretization in time of function *ϕ^v^*(*τ*), and 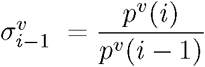 represents the proportion of infectious individuals with age of infection *i* − 1 that are still infectious on the next day. Spatially and variant explicit ℛ may be computed by solving the following system of equations:

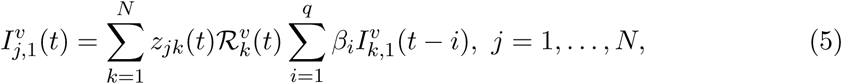

where *β_i_* is a discretization of the distribution of generation times over the age of infection. Inference of terms 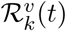 is possible via sequential Monte Carlo algorithms [16].

For the sake of completeness, let us recall the variant-implicit renewal equation i.e., when one does not consider several variants and a single process

### The multi-strain Leslie projection matrix

Let us rewrite process (4) in a matrix form and define a column vector **I**(*t*) collecting all the infectious compartments at time *t*, that is:

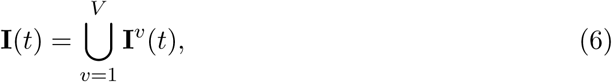

with:

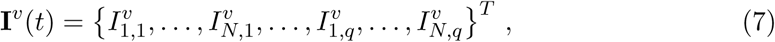

where *T* indicates matrix transposition.

The projection matrix associated with process (4) is defined such that **I***^v^*(*t* + 1) = **L***^v^*(*t*)**I***^v^*(*t*). The non-zero block **L***^v^*(*t*) for a given variant strain *v* has the structure of a so-called Leslie projection matrix and reads

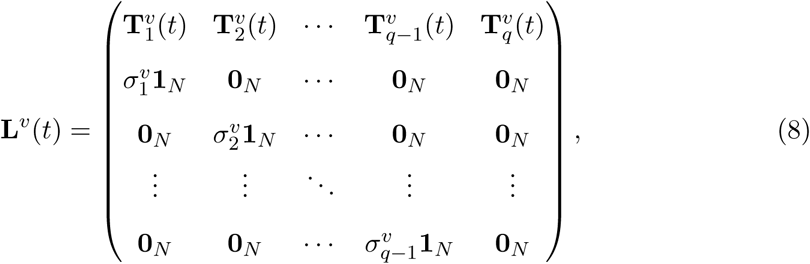

where **1***_N_* and **0***_N_* are the identity and null matrix of order *N*, respectively, and the sub-matrices 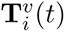 are defined as:

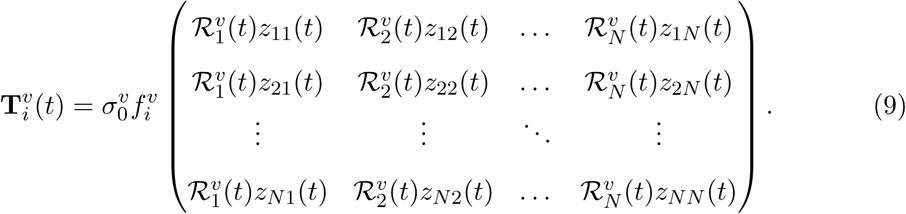

Therefore, the first *N* rows of **L***^v^*(*t*) represent the appearance of new infections. In contrast, each of the 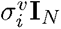 blocks corresponds to the transition of infectious individuals with age of infection *i* to their *i* + 1-th day of infection.

For multiple circulating variants, the global projection matrix **M**(*t*) is block-diagonal and reads:

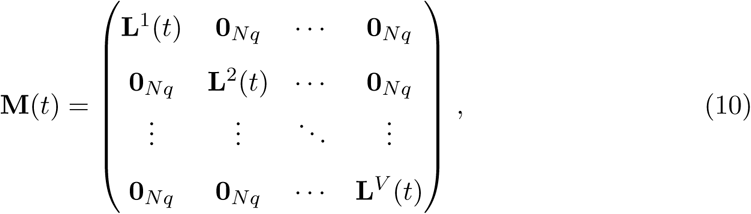

with **0***_N_q__* being a null matrix of order *N_q_*.

### Long-term analysis and the global ℛ

To assess the long-term behaviour of the system and the feasibility of a disease-free equilibrium, one first has to extract the next-generation matrix [20] and its spectral radius from the relevant Leslie projection matrix. The latter is defined as global effective reproduction number [12, 15]. To yield the next-generation matrix, each diagonal block **L***^v^*(*t*) of the general projection matrix **M**(*t*) must be split into two matrices, a transmission matrix **T***^v^*(*t*) and a transition matrix **Σ***^v^*(*t*), such that **T**^*v*^(*t*) + **Σ***^v^*(*t*) = **L***^v^*(*t*). The variant-specific next-generation matrix [21] then reads

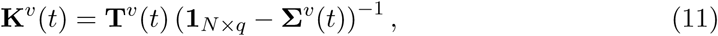

where **1***_N_q__* is the identity matrix of order *N_q_*. It is quite straightforward to prove that the following relation holds:

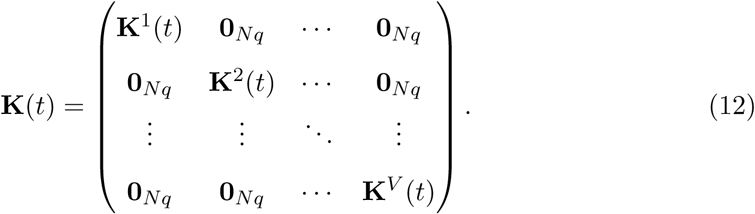

For each variant, its national 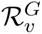 (i.e., the indicator that predicts the fate of that specific variant in the considered metapopulation) is defined as the spectral radius of the variant-specific next-generation matrix, **K**^*v*^(*t*):

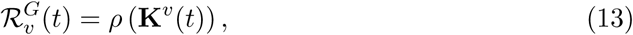

with *ρ*(·) indicating the spectral radius operator. Because of the property (12), one can easily conclude that the global ℛ*^G^*, which encompasses all variants and predicts the fate of the disease as a whole, is the largest of the variant-specific global ℛ:

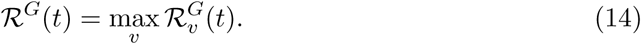

### Short-term reactivity analysis

In addition, one may analytically assess whether a short-term flare-up (which will eventually wane if the relevant global ℛ is below the unit threshold) is possible under given epidemiological conditions. Similarly to the long-term analysis, the block-diagonal nature of the Leslie projection matrix brings a substantial simplification of the relevant algebra. As such, one may define the epidemicity index for each variant subset *v* as the one-step growth of the compartments infected with variants *v*, that is:

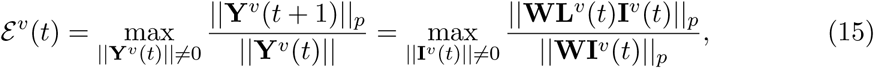

where ||*x*|| = Σ*_i_* |*x_i_*| is the *ℓ*^1^-norm of vector *x*, **I***^v^*(*t*) is the set of infectious classes (in space and age of infection) for strain *v* at time *t*, **Y***^v^* is a customary observed variable, based on **I***^v^*(*t*), and **W** is a suitable transformation matrix. Other algebraic norms (see [15]) are possible, but here, we only focus on one norm and one transformation out of simplicity. The block-diagonal property of the general projection matrix **M**(*t*), allows one to define the general epidemicity index (thus encompassing all variants) as:

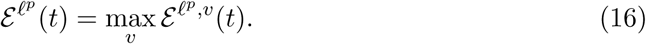

In the following, we shall discuss the short-term reactivity of the system of the observed variable **Y** defined as the vector, of length *NV* of the absolute number of infectious individuals, per variant strain and node. This vector reads

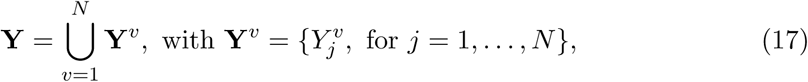

where:

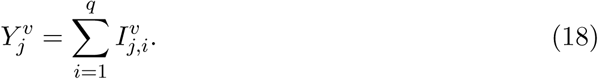

The corresponding transformation matrix **W** such that **Y**(*t*) = **WI**(*t*) takes the form:

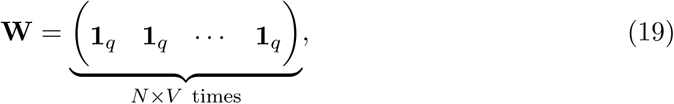

with **1***_q_* is a diagonal matrix of order *q*. Computing the epidemicity index in *ℓ*^1^-norm on this observed variable highlights the states achieving the fastest growth of one perturbation.

Given the column-stochasticity of matrix **Z**(*t*) ∀*t*, it is possible to extract a direct formula for the computation of the epidemicity index for variant *v*:

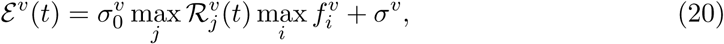

with the assumptions that 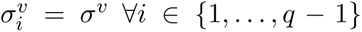, which indicate a constant rate of exit from the infectious compartments, and 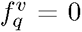, which underlines negligible infectiousness at the age of infection *τ* ≥ *q*. Reversing this formula allows one to compute the threshold reproduction number 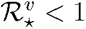 that should be achieved to guarantee that a specific variant strain is non-reactive [13]:

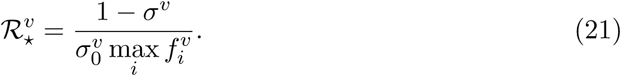

Thanks to the block-diagonal property of the Leslie projection matrix, one may easily compute the global epidemicity index as the largest variant-specific one:

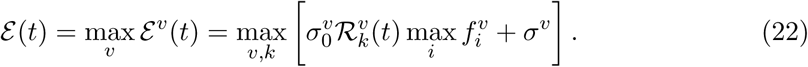

### Application to COVID-19

While this study can be implemented for any directly transmitted infectious disease, we tailor it to the COVID-19 pandemic in Italy. The distributions of generation time for the considered SARS-CoV-2 variants are assumed to be gamma-shaped, with details shown in Table 1 [22, 23, 24, 25, 26, 27]. The depletion of the infectious compartment, regardless of the considered variant, follows a negative exponential distribution, namely *p*(*τ*) = exp (−0.068*τ*), which yields *σ_j_* = exp(−0.068) ∀*j* ≥ 1, corresponding to a mean residence time within the infectious compartment of approximately 11.7 days [28]. Whenever estimates of the generation times’ distribution are not available, the serial interval is used instead, with the assumption that the incubation periods of the infector and infectee match [29, 30, 31].

**Table 1:**
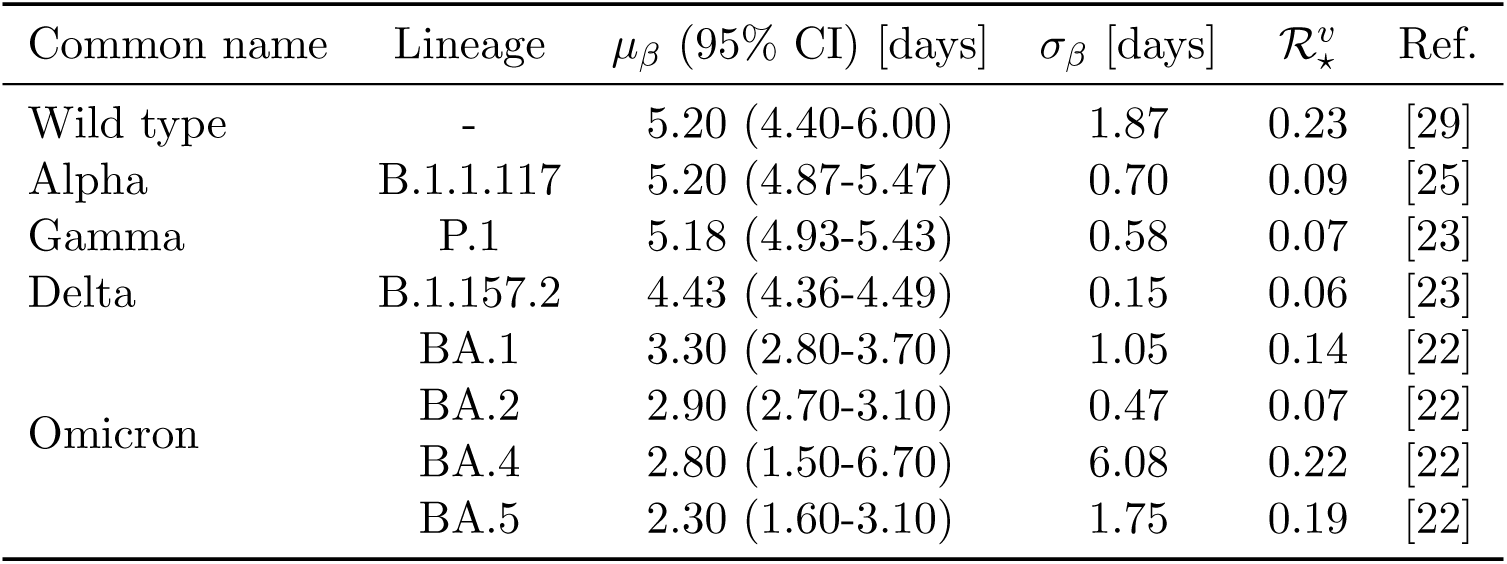
Metrics of the generation time distribution of the considered variants in the multi-variant setup. The mean of the generation time *µ_β_* is given. The standard deviation *σ_β_* is obtained from the confidence intervals. 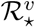 computed using Equation (21) and assuming a daily discretization of the generation time distributions are also shown.

Daily reported infections in the 20 Italian regions are gathered from the Italian Department of Public Protection (https://github.com/pcm-dpc/COVID-19). When mobility is considered, the connectivity matrix **C** is computed on the pre-pandemic connectivity matrix estimated through mobility data collected from the Italian Institute of Statistics (ISTAT) (accessible at: https://www.istat.it/it/archivio/139381) and updated during the COVID-19 pandemic via the “Workplace mobility” time series provided by the Google Community Mobility Reports (https://www.google.com/covid19/mobility/) [15]. The simulations shown here extend from February 23, 2020, through October 15, 2022, after which mobility data from the Google Community Reports were no longer available. The data on the prevalence of variants in Italy are obtained from the data portal of the European Centre for Disease Prevention and Control (ECDC), specifically from GISAID [32, 33, 34] and the ECDC’s *European Surveillance System* (TESSy). The prevalence of each variant among the sequenced samples is retained as an estimator of the prevalence of the variants among the reported infections. Only the following variants are considered for this study: the wild type (shown as “Other” in the dataset), Alpha (B.1.1.117), Gamma (P.1), Delta (B.1.157.2), and Omicron subtypes (BA.1, BA.2, BA.4, BA.5+BQ.1). All other variants subtypes are not considered as their prevalence among sequenced samples was low at all times and their inclusion would have involved an additional computational burden without producing additional information. In addition, isolated spurs of the variants occurring more than a month apart from their main transmission period are removed to reduce the computational burden. Figure 1 shows the prevalence of each variant lineage in the sequenced samples, an estimation of the prevalence of each lineage in the total reported cases of COVID-19, and a representation of the generation time distributions.

**Figure 1:**
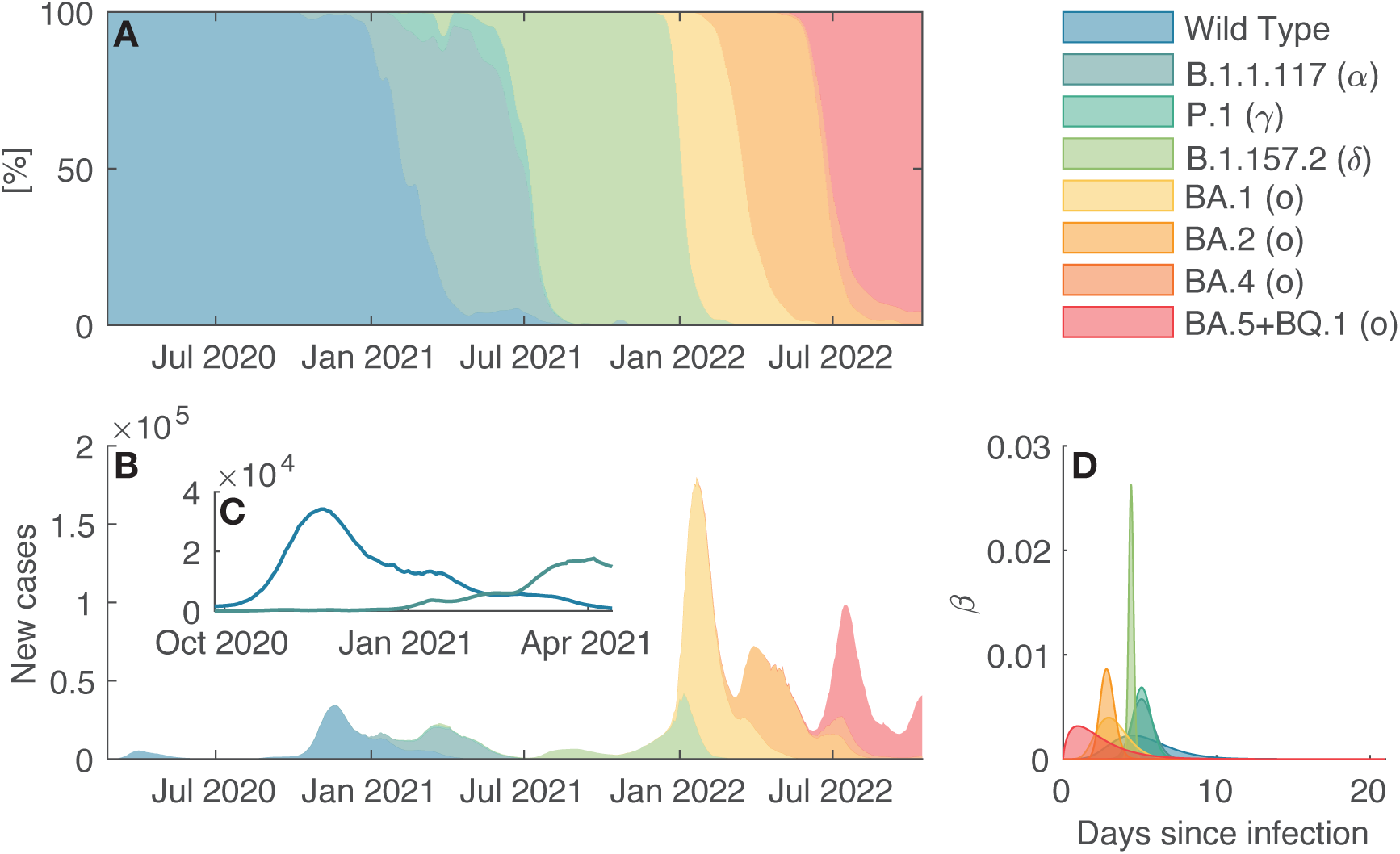
**A**:Variant prevalence for the COVID-19 pandemic in Italy; **B**: estimated number of infections attributable to each variant; **C**: same as in **B**, zoom on wild type and B.1.1.117 strains between end of September 2020 and mid-April 2021; **D**: visual representation of each variant’s generation interval distribution.

### Numerical experiments

In the following, we compare a variant-specific scenario and a non-stratified one. Concerning the non-stratified scenario, the vector **Y**_1_ has length *N*, and its transformation matrix has size (*q, N*). Process (4) loses its dependency on the variant and takes the form [15]

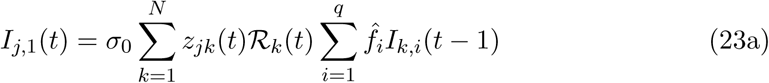

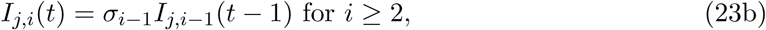

where, for the sake of precision, we compute elements 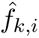 as the weighted average of the variant-specific 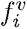, with weights equal to the active infections (per node), to allow inter-scenario comparisons:

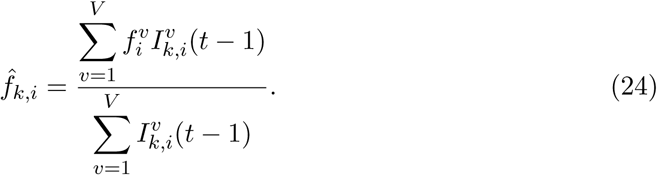

## Results

### Computation of the variant-specific effective reproduction numbers

The spatially connected effective reproduction numbers (ℛ) of the considered variant strains are computed by running a sequential Monte Carlo algorithm independently for each circulating variant [16]. We show in Figure 2 both the local variant-specific ℛ for each considered variant and, for the sake of completeness, the local variant-unstratified ℛ [15]. At the beginning of the spread of each strain, the variant-specific ℛ take a value that is substantially larger than the unit threshold—the divide between endemic spread and disease-free conditions—thus indicating a rapid initial spread, where the number of new infections caused by a given variant undergoes exponential growth. After the variant has consolidated, its ℛ then decrease to and oscillate around the unit threshold, a consequence of immunity build-up in the population (conferred by either vaccination or infection) and control measures. Interestingly, yet unsurprisingly, one may observe that, as soon as a new, fitter variant emerges, the ℛ of the previous ones tend to steadily remain below the unit threshold until the old variants disappear or become undetectable, a purely mathematical consequence of the strain waning. In addition, one may remark a larger variance among the local variant-specific ℛ for the first strains (wild type, B.1.1.117, and B.1.157.2), a trend not displayed by later strains. These differences, however, remain small, and all nodes react similarly to the circulation of SARS-CoV-2. The variant-unstratified local ℛ, on the other hand, appear much more stable. Specifically, the initial jump observed in the variant-specific is not matched by a substantial increase of the variant-unstratified above the epidemic threshold of ℛ = 1. Still, the rise in new recorded infections leads to a temporary growth of the ℛ above the epidemic threshold e.g., before January 2021, after July 2021, around January 2022 and July 2022.

**Figure 2:**
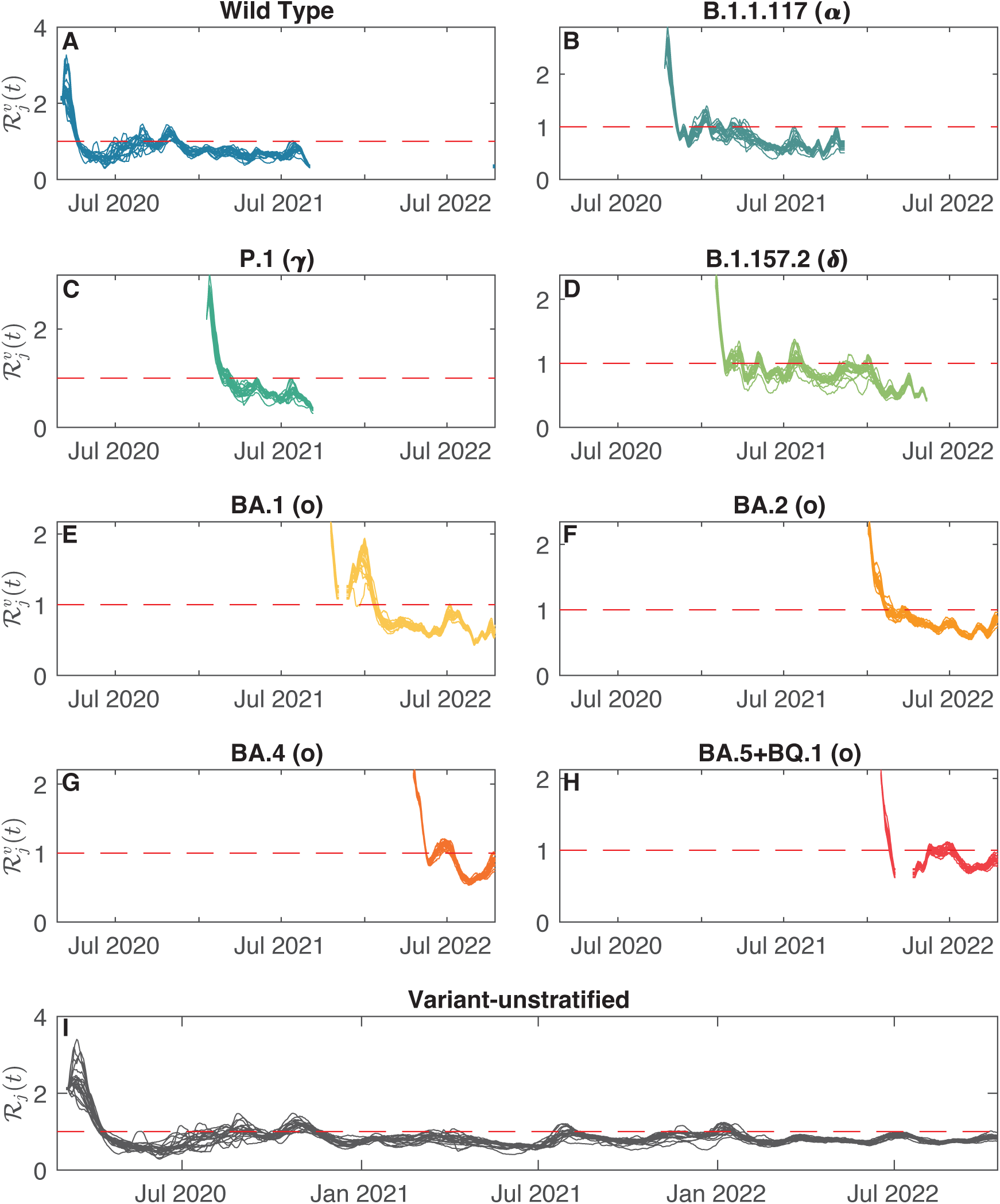
Local effective reproduction numbers for different SARS-CoV-2 strains and different model set-ups during the COVID-19 pandemic in Italy. **A**-**H**: Variant-specific ℛ; **I**: Variant-unstratified ℛ.

### Computation of the national epidemiological indices

The national ℛ*^G^* and epidemicity index *ε*(*t*), are shown in Figure 3 for both the variant-specific and variant-unstratified scenarios. As previously observed (Figure 2), whenever a new variant emerges, its local variant-specific ℛ are characterized by an initial spike. This sudden jump is also seen in the global variant-specific ℛ*^G^* is computed, essentially the largest value among the national variant-specific ℛ (Figure 3A).

**Figure 3:**
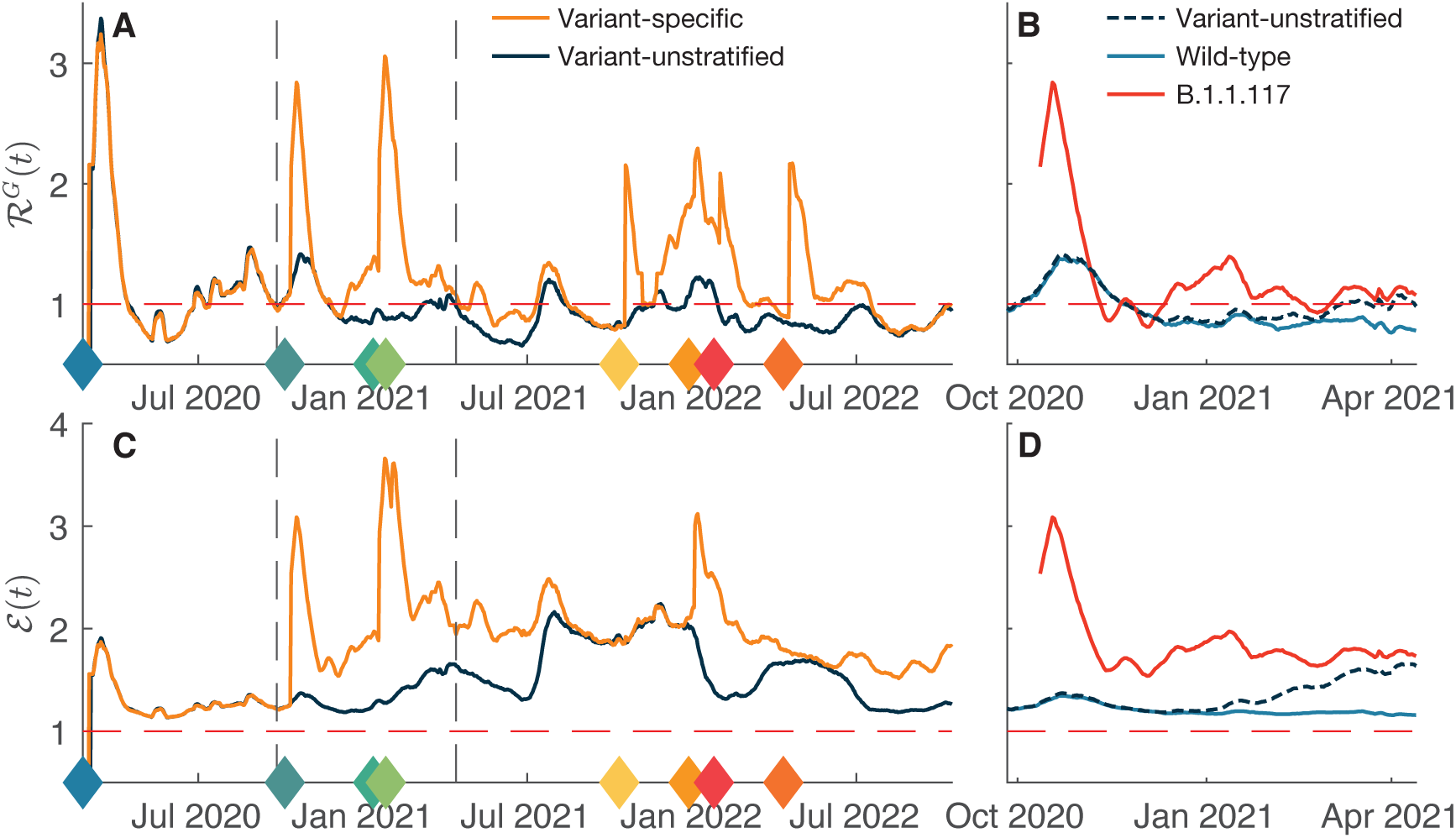
General epidemiological indices evaluated for different model set-ups. **A**: national effective reproduction number, dashed vertical lines highlight the zoom shown in panel **B**, filled diamonds mark the emergence of a new variant and are colour-coded according to Figure 1; **B**: as in **A**, showing national ℛ for wild type and B.1.1.117 (here highlighted in red) strains between end of September 2020 and mid-April 2021 against the variant-unstratified national ℛ. **C**: epidemicity index; **D**: as in **B**, but showing the epidemicity index.

The emergence of a new variant often brings the two time series (variant-specific and -unstratified global reproduction numbers) to the two different sides of the unit threshold, the variant-unstratified denoting disease-free equilibrium conditions. This is a direct consequence of the fact that, while the variant-unstratified ℛ*^G^* only focuses on the total number of recorded infections, the variant-specific one detects whether any specific strain is growing. To better disentangle the behaviour of the variant-specific ℛ*^G^*, we investigated the period during which the B.1.1.117 (Alpha) variant has replaced the wild type (see Figure 3B) by computing the national ℛ for both strains. At the emergence of the Alpha variant in October 2020, the new infections caused by this strain followed a steady exponential growth. Nonetheless, these additional infections did not shift the total number of recorded infections until January 2021 (Figure 1C). This is reflected in the national ℛ of the Alpha variant, which immediately diverged from the variant-unstratified national ℛ, which remained closer (and below the epidemic threshold) to the national ℛ of the wild type, a waning strain, and only started growing above the epidemic threshold around March 2021.

One may observe larger increases for the first new variants (Alpha and Delta), at which emergence the ℛ*^G^* skyrockets to values up to three times higher. Conversely, later variants are characterized by minor, but still relevant spikes. The variant-unstratified ℛ*^G^* does not display this behaviour and stabilizes around the epidemic threshold with only modest increases, a trend also shown by the local variant-unstratified ℛ. The two time series (variant-specific and unstratified), however, converge as soon as a variant establishes as the only circulating strain, as seen during the first phase of the epidemic up to October 2020 and, briefly, around autumn 2021.

The epidemicity index (Figure 3C) also holds a similar behaviour, with sudden spikes at the emergence of a new variant. However, the divergence between the variant-specific epidemicity index and the variant-unstratified one appears wider than the discrepancy observed for the ℛ. This is apparent throughout the investigated timespan, and even more so during the replacement of the wild type with the Alpha variant (Figure 3D). Once again, the gap between the epidemicity index of the newer strain and that of the one being replaced proves wider than the difference between the national reproduction numbers of the two variants. A higher epidemicity index of the Alpha strain is a consequence of a higher ℛ (Figure 3B) and a lower epidemic threshold (see Table 1). Owing to a different generation time’s distribution, the threshold reproduction number 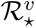 of the Alpha variant is much lower than that of the wild type, meaning that control measure should target a much lower ℛ to achieve unreactive conditions.

## Discussion

The arrival of new pathogenic variants during an ongoing epidemic induces shifts in the generation time’s distributions and basic reproduction numbers, which are not accounted for in current algorithms for the computation of epidemiological indices. Using an adequately modified existing sequential Monte Carlo algorithm, we included the pathogen’s stratification into several variants and updated the epidemiological indicators. Our main result shows that adding a stratification in variants leads to substantially higher values of the reproduction number and epidemicity index, with relevant implications for epidemic control.

First, we computed the variant-specific local ℛ via already validated techniques [16]. Our computations show that the ℛ of an emerging variant takes values well above the epidemic threshold, whilst both ℛ of the circulating strain and the variant-unstratified one merely fluctuate around or below said threshold. This is normal behaviour for the strain being replaced, as its disappearance is directly highlighted by a ℛ *<* 1 for a prolonged period. On the other hand, the behaviour of the variant-unstratified ℛ only focuses on the total number of recorded infections and is therefore insensitive as to whether one strain has begun growing until the emergent strain overcomes the previously circulating ones and drives the total number of infections up itself.

This behaviour also emerged with the global ℛ*^G^*, which accounts for all variants spreading in a connected metapopulation. While the variant-unstratified ℛ*^G^* showcases the same behaviour of the local ℛ, being only dependent on the total number of infections, the variant-specific one displays sudden spikes at the appearance of a newer and evolutionary fitter strain. These jumps underline the presence of a variant growing much more rapidly than the total number of infections, as was the case around November 2020 when the Alpha strain started replacing the wild type. This replacement went unnoticed in terms of the effective reproduction number until March 2021, when Alpha became the dominant strain and the variant-unstratified ℛ*^G^* finally started growing above the epidemic threshold. The variant-specific ℛ*^G^* anticipates this growth and highlights a potentially concerning situation well before a dominant strain is replaced. Failing to consider variants’ stratification may thus lead to an underestimation of the severity of an epidemic situation.

The epidemicity index follows a qualitatively similar trend, and the variant-specific time series also diverges from the variant-unstratified at the emergence of a newer and fitter strain. Similar spikes may be observed throughout the considered period. In addition to such spikes, the divergence between the variant-specific and variant-unstratified time series appears substantially larger than the one observed with the reproduction numbers. This is a direct consequence of the higher reactivity of several strains than that of the wild type, as characterized by lower ℛ^⋆^ (Table 1). For this reason, epidemic control should not simply aim at achieving ℛ^*G*^ < 1, but rather ℛ^*G*^ < ℛ^⋆^.

Our illustrative experiments have two main limitations. Firstly, to implement the framework introduced here, one has to gather or analytically derive the number of new infections caused by each relevant strain. While in practice, the chance for a variant to be sampled might be slightly different from its actual prevalence among the sequenced samples, this probability is unlikely to swiftly change over time and impact the computation of ℛ. Secondly, our framework requires the variants’ generation time distributions. This might prove challenging at the emergence of a new variant (or at the emergence of the wild type of the pathogen itself) as estimations of the serial interval and the generation time’s distribution usually require a few weeks of observations to be estimated. However, implementing even early estimates of the relevant distributions still helps understand a changing situation. Furthermore, strain replacement, during which the variant-specific frameworks give substantially different results than the variant-unstratified one, occurs in a few months.

In conclusion, our results show that whenever variant stratification is considered, embedding the pathogen’s stratification in variants in algorithms for computing the aforesaid epidemiological indices produces substantially different estimates of the epidemiological indicators, with wide ramifications for control measures. Our updated framework detects whether one specific strain is growing more rapidly than the total number of infections and, therefore, anticipates a possible growth of new infections once the emerging variant becomes the dominant strain. Because our framework combines stratification in both space and variants, it provides more precise estimates of the epidemiological metrics in a comprehensive environment. This is particularly relevant when the problem involves several communities and co-circulating strains. Finally, the sequential Monte Carlo process makes it possible to update the effective reproduction number and epidemicity index estimates as new surveillance data becomes available, with great practicality.

## Data availability statement

All codes and data involved in this study are available at the following public GitHub repository: cristianotrevisin/epidemiological-indices-metapopulation.

## Declaration

All authors declare no competing interests.

## Authors contributions

Conceptualization: C.T., A.R.; Data Curation: C.T., L.M.; Formal Analysis: C.T., L.M., M.G., V.C., A.R.; Funding Acquisition: A.R.; Investigation: C.T., L.M., M.G., V.C., A.R.; Methodology: C.T., M.G.; Software: C.T.; Validation: C.T., L.M., M.G., V.C.; Visualization: C.T., L.M., V.C.; Writing – Original Draft Preparation: C.T.; Writing – Review & Editing: C.T., L.M., M.G., V.C., A.R.

## Acknowledgement

C.T. and A.R. acknowledge funding from the Swiss National Science Foundation via the project ‘Optimal control of intervention strategies for waterborne disease epidemics’ (grant number 200021-172578). L.M. acknowledges funding from the Italian Ministry of University and Research through the project ‘Epidemiological data assimilation and optimal control for short-term forecasting and emergency management of COVID-19 in Italy’ (FISR 2020IP 04249).

